# Data extraction error in pharmaceutical versus non-pharmaceutical interventions for evidence synthesis: Study protocol for a crossover trial

**DOI:** 10.1101/2022.08.27.22279301

**Authors:** Yi Zhu, Pengwei Ren, Suhail A.R. Doi, Luis Furuya-Kanamori, Lifeng Lin, Xiaoqin Zhou, Fangbiao Tao, Chang Xu

## Abstract

**Background:** Data extraction is the foundation for research synthesis evidence, and it is often time- and labor-consuming, and prone to errors. Whilst data extraction errors frequently occur in the literature, an interesting phenomenon was observed that data extraction error tend to be more common in trials of pharmaceutical interventions compared to non-pharmaceutical ones. This phenomenon has not been verified by high-quality evidence, the elucidation of which would have implications for guidelines, practice, and policy.

**Methods and analyses:** We propose a crossover, multicenter, investigator-blinded, trial to elucidate the potential variants on the data extraction error rates of meta-analyses with pharmaceutical against non-pharmaceutical interventions. Eligible 90 participants would be 2^nd^ year or above post-graduate students (e.g., masters, doctoral program). Participants will be randomized to one of the two groups to complete pre-defined data extraction tasks: 1) group A will contain 10 randomized controlled trials (RCTs) of pharmaceutical interventions; 2) group B will contain 10 RCTs of non-pharmaceutical interventions. Participants would then be assigned to the alternative group for another round of data extraction, after a 30 mins washout period. Finally, those participants assigned to A or B group will be further 1:1 randomly matched based on a random-sequenced number, for the double-checking process on the extracted data. The primary outcome will be the data extract error rates of pharmaceutical intervention group and non-pharmaceutical group, *before* the double-checking process, in terms of the cell level, study level, and participant level. The secondary outcome will be the data error error rates of pharmaceutical intervention group and non-pharmaceutical group, *after* the double-checking process, again, in terms of the cell level, study level, and participant level. Generalized linear mixed effects model (based on the above three levels) will be used to estimate the potential differences in the error rates, with a log link function for binomial data. Subgroup analyses will account for the following factors i.e., the experience of individuals on systematic reviews, and the time used for the data extraction.

**Ethics and dissemination:** This study has been approved by the institutional review board of Anhui Medical University (No.83220405). Findings of the study will be presented at international scientific meetings, and publish in peer-reviewed academic journal.

**Trial registration:** Chinese Clinical Trial Register Center (Identifier: ChiCTR2200062206).

**Strengths and limitations of the study:**

- This will be the first trial to compare data extraction error rates in pharmaceutical intervention and non-pharmaceutical intervention studies for research synthesis.
- This will be the third randomized trial on the strategy of data extraction in the world and the first in the Asia-Pacific region.
- The use of a crossover design provides a valid way to reduce the potential impact of heterogeneous contexts of the studies and thus is expected to provide robust evidence to support better evidence synthesis practice.
- We will restrict the participants to 2^nd^ year post-graduate students or above to ensure the feasibility of the trial; this restriction will no doubt impact the representativeness of the samples.
- A group of useful strategies (eg. use U disk and isolate signal etc.) should be taken to minimize the impact of the possible sharing of the completed extraction table among the participants.

## Introduction

In an era of evidence-based medicine, research synthesis is the backbone of healthcare practice and it governs the guideline-developing, decision-making, as well as policy-formulating [1]. Systematic review and meta-analysis has been one of the most important sources of evidence and thus the validity of such evidence directly determines the reliability and quality of healthcare administration [2]. Unfortunately, in real-world, the evidence generated from systematic reviews and meta-analyses is far from valid or trust-worthy due to a multiple reasons, where one of which would be errors during data extraction — as recorded in previous studies, as much as 85% of the systematic reviews faces serious issue in data reproducibility [3-7].

Data extraction is a crucial step for any type of evidence synthesis; it undertakes the important role of information transformation, from one ‘node’ to another. This means any error during this process would inevitably distort the original information and thus may bias the final evidence. In our recent study, in 288 meta-analyses with data extraction errors, 12.8% (n = 39) were moderately or largely impacted in terms of the magnitude of the effects [7]. The Cochrane Collaboration has highlighted the importance of qualified data extraction for informed decision-making and recommended the application of the duplicate data extraction strategy in Cochrane reviews [8]. The AMSTAR (Assessing the Methodological Quality of Systematic Reviews), a popular cheklist for research synthesis, also highlights the importance of good data extraction practice [9].

While evidence from two randomized trials suggest some benefits of duplicate data extraction for improving data reproducibility [10, 11]; our empirical investigation suggest that the error rates may differ by the type of interventions. We found that meta-analyses assessing pharmaceutical interventions had almost twice the error rate than investigating non-pharmaceutical interventions [6,7]. The above findings sparked our interest in why the error rates differ in meta-analyses with pharmaceutical and non-pharmaceutical interventions, and whether duplicate data extraction could ‘trade-off’ such preference on the occurrence of errors?

Therefore, in this protocol, we described a planned crossover, multicenter, investigator-blinded trial with the aim of elucidating the aforementioned questions.

## Methods

### Ethics and trial registration

This study has been approved by the institutional review board of Anhui Medical University (No.83220405), and has been registered at the Chinese Clinical Trial Register Center (Identifier: ChiCTR2200062206). The study is designed in line with the CONSORT statement [12], and the reporting of the current protocol follows the SPIRIT (Standard Protocol Items: Guidelines for Interventional Trials) 2013 checklist [13].

### Study design and settings

This is a 1: 1 designed, randomized, multicenter, investigator-blinded, cross-over trial. The trial will be conducted in three centers in China, i.e., Anhui Medical University (leading medical university in Anhui province), Taihe Hospital (leading hospital in Shiyan city and the top 10 hospitals in Hubei province), and Guizhou Provincial People’s Hospital (one of the three leading hospitals in Guizhou province). All three centers have ongoing research and teaching programs of evidence-based medicine for post-graduate students. We have prepared a appropriate classroom in three centers. Participants will be required to bring laptop to the classroom for data extraction. Some investigator will help and supervise the participants.

The trial will involve three stages of data extraction (Figure 1). In the first stage, One of the two data extract groups based on crossover design will extract studies data of pharmaceutical interventions first, and then extract data of non-pharmaceutical interventions. Similarly, the other group will perform the same steps in the reverse order, i.e., extract data of non-pharmaceutical interventions first, and then extract data of pharmaceutical interventions. Due to blinding, we do not know which group was performed in which arrangement. Participants will be randomly allocated into one of the two groups for data extraction that with studies of pharmaceutical interventions or with studies of non-pharmaceutical interventions that will be prepared in advance. In the second stage, after a washout (i.e., break) period of 30 minutes, participants who performed data extraction on studies of pharmaceutical interventions will be switched to non-pharmaceutical interventions for another round of data extraction, and vice versa.In the third stage, participants in of the two groups assigned in the first period will be randomly matched (1:1) with ones from diverse groups among these based on a random-sequenced number, for the double-checking process on the extracted data. For example, If participant A and participant B in diverse groups are assigned to the same random-sequenced number, they would be matched to form a pair. Without the same number would not be allowed to participate in the double-checking process. In each pair, the previously extracted results will be checked by the two participants. After the discussion and agreement, the final results that the participants ponder will be filled in a new sheet, and the inconsistent places will be marked (the number color turns red). During each of the three stages, participants will be granted at least 30 mins break before they enter the next period.

**Figure 1.**
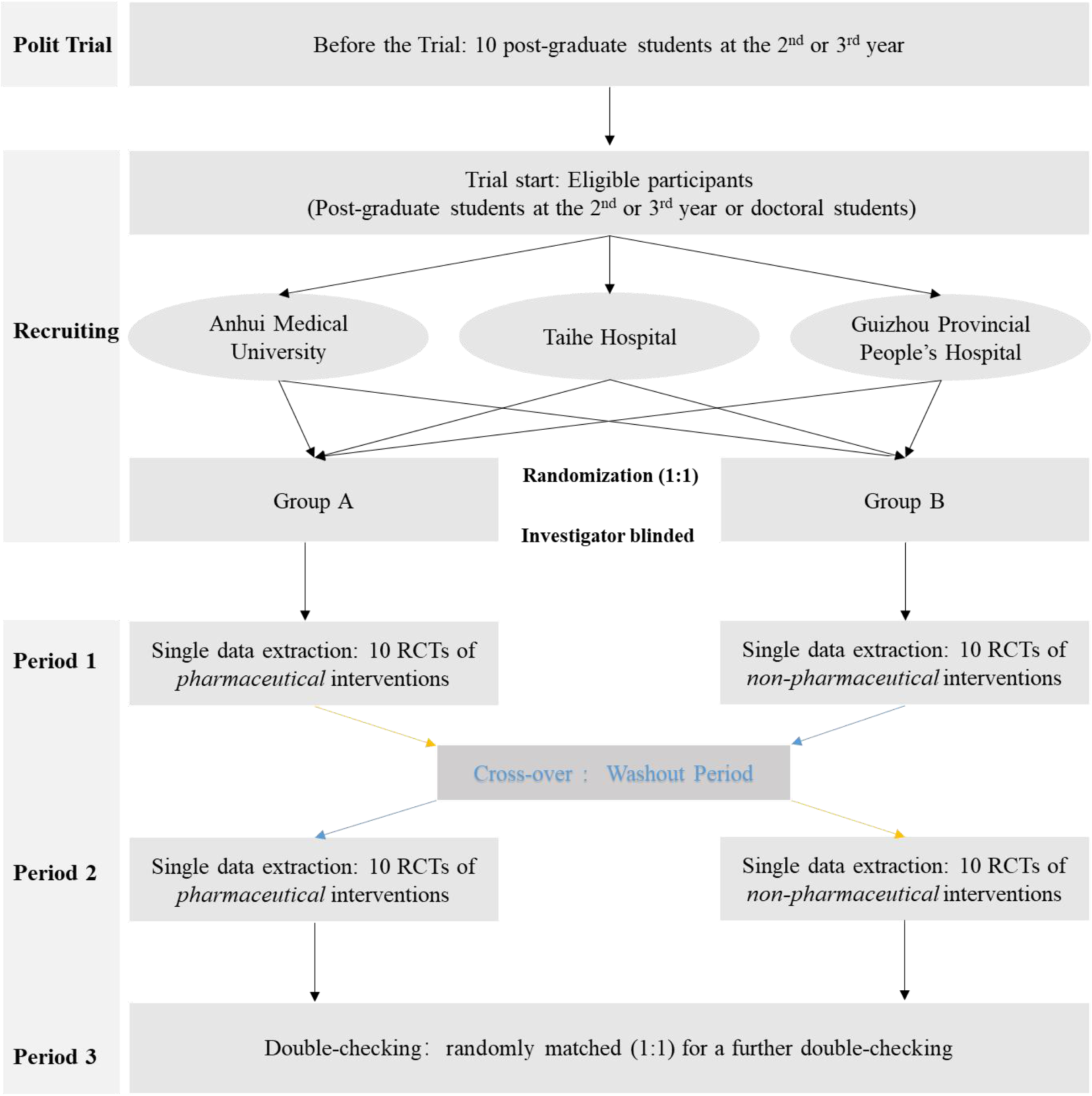
Study design.

The studies used for data extraction in the two groups will be identified before the start of the trial, based on our previous well-established database of meta-analyses of adverse events (binary outcomes) [7]. The database covers 201 systematic reviews of randomized controlled trials with 829 meta-analyses with pharmaceutical or non-pharmaceutical interventions, and all the meta-analytical data in the database have been carefully checked for their validity. The lead investigator (CX) will select one meta-analysis of pharmaceutical intervention and one of non-pharmaceutical intervention and remove the existing meta-analytic data to form an spreadsheet with only the study list and necessarily column titles (but without data) as the template for data extraction (Table 1). By reviewing previous meta-epidemiological studies, we set the number of studies for data extraction in each group to 10 [14, 15]; therefore, each of the selected meta-analyses should contain at least 10 studies. If the number of included studies exceeds 10 in the selected meta-analysis, a simple random sampling scheme will be employed to randomly select 10 studies.

**Table 1.** Example of the data extraction form.

| Section 1: Basic Information |  |  |  |  |  |  |  |
| --- | --- | --- | --- | --- | --- | --- | --- |
| Student ID |  |  | Age |  |  | Gender |  |
| Background |  |  | Experience in publication |  |  | Experience in Meta-analysis |  |
| Section 2: Key Components of PICO |  |  |  |  |  |  |  |
| Population | Any population | Intervention | Amoxicillin | Comparison | Placebo | Outcome | Diarrhea |
| Section 3: Data Extraction form |  |  |  |  |  |  |  |
| Study | Intervention (Amoxicillin) |  |  | Comparison (Placebo) |  |  | Electronic timer |
|  | Cases | Total | Treatment Duration (wk) | Cases | Total | Treatment Duration (wk) |  |
| Example | 3 | 8 | 16 | 1 | 8 | 16 | Click to Start |
| Burke et al |  |  |  |  |  |  |  |
| Jørgensen et al |  |  |  |  |  |  |  |
| Meltzer et al |  |  |  |  |  |  |  |
| Merenstein et al |  |  |  |  |  |  |  |
| Nduba et al |  |  |  |  |  |  |  |
| Taylor et al |  |  |  |  |  |  |  |
| Wald et al |  |  |  |  |  |  |  |
| Baecher et al |  |  |  |  |  |  |  |
| Esposito et al |  |  |  |  |  |  |  |
| Trehan et al |  |  |  |  |  |  |  |

### Participants, intervention and control group

Individuals with medical or health science backgrounds that are learning systematic reviews, preparing an ongoing systematic review, or already have experience in conducting systematic reviews are eligible to participate. This may include clinicians, nurses, healthcare policy makers, medical scientists, and medical students. Whereas English is not the native language of Chinese that individuals without qualified English reading skills may have poor performance in data extraction, and the fact that medical students played the main role in data extraction in the majority of the published systematic reviews, we will only consider students at the 2^nd^ year of their post-graduate program and above (e.g., doctoral program). Based on a pilot study findings, we expect the time for data extraction for all the three stages to be between 3 and 5 hours. Participants will be compensated with 150 RMB (about 22 USD, 4.5 to 7.5 USD/hour) for their time.

The primary aim of the trial is to examine the error rates of data extraction in RCTs of pharmaceutical over non-pharmaceutical interventions and the role of duplicate extraction in reducing error rates in evidence synthesis practice. Thus, the intervention of the current trial is the double-checking scheme in the third period of the trial. While for the control, based on the design of this trial, two controls will be involved. The first control is single data extraction, namely, data extraction with only one individual (first and second period), without the involvement of any other individuals. For the single data extraction, self-checking is allowed. The second control is the non-pharmaceutical RCTs, compared with the pharmaceutical RCTs, in terms of the data extraction error.

### Randomization, blinding, and allocation concealment

A third party, a doctor proficient in randomization and not involved in the trial, will generate the random sequence using a computer random-number generator right after the enrollment. Participants will take part in the baseline evaluation (e.g., age, expertise, experience in systematic reviews). After providing written informed consent, if eligible participant accepted, then he/she is enrolled. Then, participants will be randomized to one of the two groups with a 1:1 ratio through simple randomization. The random sequence will be sent to the participants directly by the third party through email 15 mins before the formal trial separately, with corresponding data extraction form and PDF files of related 10 RCTs of the referred group.

Investigators will be blinded in the whole process owing to the aforementioned process (the investigators will not know the random sequence until the third party unblind the sequence). However, it will not be possible to blind the participant because participants will receive the data extraction form and the affiliated materials, they will know the intervention type of the RCTs they are about to perform the data extraction. Allocation will be concealed through a unique password-protected data extraction form which will be allocated directly by the third party. In addition, outcome accessors and statistical analysts will be blinded owing to the employment of a third party. To prevent potential exchange on the extracted data among participants within or between groups, the 10 studies in the data extraction form will be ranked randomly for each participant.

### Outcomes

The primary outcome will be the error rates on data extraction of pharmaceutical intervention group and non-pharmaceutical group, *before* the double-checking process, in terms of the cell level, study level, and participant level. The secondary outcome will be the error rates on data extraction of the pharmaceutical intervention group and the non-pharmaceutical group, *after* the double-checking process, again, in terms of the cell level, study level, and participant level.

In addition, the time-standardized error rates of pharmaceutical over non-pharmaceutical intervention before and after the double-checking process are also of interest. For each individual, the time taken to complete the data extraction in each period is defined as the sum of the time spent on each of the RCTs. This will be recorded through a self-programmed Excel micro by the third party. In brief, when the participants click the ‘Start’ button, the program will start to record the time; and when they click the ‘Stop’ button, the program will stop recording the time. Participants may leave the room for some private reasons that could prolong the time as long as they forget to click ‘stop’. To avoid such an overestimation, any participants will be asked to inform the investigators if they need a short leaving so that the investigators can record the time in leaving of the participants, and this will be subtracted from the total time.

### Follow-up

The follow-up of the trial would be from the beginning of the data extraction to the complementation. No further follow-up will be required due to the aim of the trial. As a result, we expect a low dropout rate.

### Sample size calculation

We used the following formula [16] to estimate the sample size requirements for each of the two groups for an equivalence test:

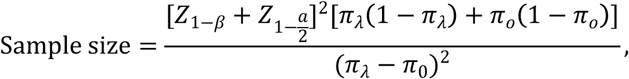

where *Z* is the standard score that refers to the number of standard deviations from the mean, *a* is the significance level, and *β* is the statistical power that reflects the ability to reject the null hypothesis when there is a true effect. In addition, *π*_*λ*_ is the error rate of data extraction in intervention arm and *π*_*o*_ is the error rate in control arm. Here, the sample size is estimated in terms of both study level and participant level. For the study level, the total sample size means the number of participants (*n*) multiple the number of studies (*k*), namely, *n* ∗ *k*; for the participant level, the total sample size means the number of participants (*n*).

For both levels, we considered an *a*=0.05 and *β*=0.8, while the event rates *π* differ. For the study level, based on the previous two trials [10,11], *π*_λ_ is expected to be between 15.41% and 19.90%, while *π*_0_ is about 8.87% to 10.20%. For the participant level, based on our empirical investigation and other studies [6, 7], about 65% of the meta-analyses had data extraction errors — some referred to pharmaceutical interventions, and some referred to non-pharmaceutical interventions, indicating that *π*_λ_ would be greater than 65%, while *π*_0_ would be less than 65%. Therefore, we empirically set *π*_λ_ = 20% for study level and 80% for participant level, while *π*_0_ = 10%for study level and 50% for participant level. Based on such settings, we obtained a sample size for each group of 12 (118/10) in terms of the study level and 40 (398/10) in terms of the participant level. Under a dropout rate of 10%, we take 45 as the minimal sample size of each group, and thus at least 90 participants are needed in total.

### Participants’ recruitment

Investigators will paste advertisement poster in the main buildings (i.e., teaching building, dining hall) of the three centers to make sure the majority of students can reach out for the recruiting information. To maximize the visibility of the advertisement, investigators and their colleagues will share the e-poster on their own social media (e.g., WeChat) or in community groups. Subjects who are willing to participate will also be encouraged to invite their friends to join this study. The recruitment started on 25 August 2022.

### Data collection

Data collection will be done along with the data extraction form mentioned above. The following baseline information will be collected from participants: age, gender,, experience in systematic reviews, and experience of publication. For the sake of privacy, the name of participants will not be collected; instead, the student identifiers for each participant will be collected. Any additional information that is considered useful at any period of the trial will be collected through a face-to-face interview. Time information on data extraction will be automatically recorded by the macro as aforementioned.

### Statistical analyses

Baseline characteristics will be summarised by the data type. For discrete variables (e.g., gender, professional background, experience on systematic reviews), frequency and proportion will be summarized, and for continuous variables (e.g., age), the mean and standard deviation (SD) or median and interquartile range (IQR) will be presented. Baseline demographic data will be compared using an independent-sample *t*-test or chi-squared test as a verification of the implementation of the randomization process.

For the main analysis, both intention-to-treat and per protocol principles will be used to examine the potential difference in data extraction error rates amongst the above comparisons. Considering that the estimation of participant-level error rate would be impacted by the study level and the cell level estimates, we will establish a generalized linear mixed model by treating each cell as level 1, study as level 2 and participant as level 3 to address this problem. The risk ratio (RR) will be used as an effect estimator under the binomial distribution with a log link faction [17, 18]. While time used for data extraction is expected to impact the error rate, the time-standardized rate measured by incidence risk ratio (IRR) will be estimated under the mixed Poisson model [19].

Subgroup analysis will be employed for the following factors that may impact the quality of data extraction, including the gender, experience of individuals on systematic reviews, time used for the data extraction, and experience on publication of the participants. All the analyses will be conducted using Stata/SE 16.0 (Stata Crop LCC, College Station, TX), with a significance level of 0.05. The statistical analysis will be done by a statistician who will be blinded to the allocation information.

## Discussion

In this protocol, we describe the design, implementation, and analysis plan of a forthcoming trial to establish informed evidence for qualified data extraction in evidence synthesis practice. To the best of our knowledge, this will be the first trial that compares the error rates of data extraction by type of intervention. The study will be the third randomized trial on the strategy of data extraction, while the first in the Asia-Pacific region. The study will also provide evidence on the potential benefits of duplicate data extraction on data reproducibility, in reducing errors and trading-off the potential negative impacts of intervention type on errors. Through a crossover design, the study presents a valid approach to reduce the potential impact of the heterogeneous contexts of the studies and thus is expected to provide robust evidence to support better evidence synthesis practice.

There are some limitations in terms of the design and implementation. First, to ensure the feasibility of the trial, we restrict the participants to 2^nd^ year post-graduate students or above; this would no doubt impact the representativeness of the samples. Indeed, clinicians are one of the key contributors to published systematic reviews and meta-analyses, who will not be represented in this trial. Second, we will use a spreadsheet for participants to perform the data extraction. Although the spreadsheet will be encrypted and protected, there still be risk that participants could share the data with other participants — this would ‘contaminate’ the dataset and introduce bias.

Fortunately, some useful strategies are taken to prevent this case from happening, for example, conducting a pilot trial to evaluate the risk, disrupting the order of the study list in the sheet, removing duplicated data sheet if the information is identified as totally the same by different participants. The random assignment and blinding scheme would also be a valid approach to present the case.

In summary, the conduction of this trial is expected to provide useful evidence to guide the data extraction practice for further systematic reviews authors and new evidence for methodologist to design a better data extraction strategy.

## Data Availability

The data will be shared with the public after the publication of the trial.

## Declarations

### Ethics

This RCT was registered on the Chinese Clinical Trial Registry (Identifier: ChiCTR2200062206). Ethics approval was obtained from the Anhui Medical University IRB (No.83220405).

### Dissemination

Results of this protocol will be published in a peer-reviewed journal, and disseminated at local, national and international meetings. The paper describing the key findings will be submitted to a journal within 12 months of the trial completion. Participants will not be identified in any publications or presentations resulting from this study.

### Data sharing

The data will be shared with the public after the publication of the trial.

